# Heart failure grading using single-lead electrocardiography

**DOI:** 10.1101/2020.10.08.20209700

**Authors:** Eriko Hasumi, Katsuhito Fujiu, Ying Chen, Yu Shimizu, Tsukasa Oshima, Hiroshi Matsunaga, Jun Matsuda, Takumi J Matsubara, Nobuaki Fukuma, Liu Yuxiang, Junichi Sugita, Yukiteru Nakayama, Akiko Saga, Gaku Oguri, Toshiya Kojima, Yujin Maru, Morio Shoda, Issei Komuro

## Abstract

Convolutional neural networks (CNNs) applied to electrocardiograms (ECGs) have been showing utility for detecting left ventricular (LV) dysfunction^1^. Although early detection of reduced LV ejection fraction (rEF) could improve handling of heart failure (HF) with rEF (HFrEF), it is not sufficient to detect HF with preserved EF (HFpEF). Here we developed a CNN algorithm to classify the severity of HF based on single-lead ECG data, irrespective of EF. We trained a CNN using ECG data and the HF classification from 7,865 patients with HF. The CNN achieved an area under the receiver-operating characteristic curve (AUC) of 0.996 for distinguishing patients with HF of various severity from healthy controls. It is anticipated that early detection of HF and therapeutic management of HF patients can be improved by employing this CNN with a single-lead ECG device.

## Main

An estimated 64.3 million people worldwide are living with HF^2^. Moreover, because of population aging, global population growth, and improved survival after diagnosis, the total number of HF patients is rising^3^. Currently, patients are categorized as having HF with reduced ejection fraction (HFrEF: left ventricular ejection fraction (LVEF) <40%), mid-range EF (HFmrEF: LVEF 40-49%), or preserved EF (HFpEF: LVEF ≥50%)^4^. In recent studies, CNN algorithms for screening low LVEF using features of the ECG were developed and confirmed to exhibit favorable performance^1^. Attia et al. reported that their AI algorithm provided a high degree of discrimination between EF≤35% and EF>35% based on 12-lead ECG (AUC 0.93)^1^. Cho et al. also reported an AI algorithm that detected low LVEF, defined as an EF<40% (AUC 0.961 for 12 lead-ECG and AUC 0.929 for single-lead ECG), with reasonable accuracy^5^. However, the LVEF-based taxonomy of HF and its implications have been criticized for leading to oversimplification of this complex syndrome^6^, as there is an unexpectedly high rate of adverse outcomes among patients with HFpEF^7^.

Prevention of progression and recurrent hospital admissions in patients with established HF continues to be a challenge^8^. In particular, the incidence of HFpEF is rapidly rising such that it is becoming the most common form of HF^9^. One in five inpatients with HFpEF were readmitted within 30 days of hospital discharge, and more than half were readmitted within one year10 Echocardiography is central to the diagnosis of HFpEF, which requires detection of structural abnormalities, such as left ventricular hypertrophy or left atrial enlargement, or signs of diastolic dysfunction^11^. However, because the number of times an echocardiograms can be repeated is limited, they should only be done in specific situations^8^.

The New York Heart Association (NYHA) classification is a patient symptom-based HF classification system that is essential to the current therapeutic perspective^12^. A HF patient’s symptoms can be used to classify the severity their HF and to monitor the effects of therapy, and they are more closely related to a poor prognosis if a high score persists after therapy^13^. Moreover, the NYHA classification could be applied to both the HFrEF and HFpEF categories if it were easy to assess the category daily from HF patients after their discharge from the hospital, which could enable early detection of HF recurrence and early intervention.

However, association of NYHA classifications with HF symptoms by the patients themselves or non-cardiologists may be difficult^14^. We therefore developed a CNN for automatic classification of HF patients according to the NYHA classification using only lead I of an ECG. At present, acquiring a lead I ECG at home is possible with numerous portable devices. Here we show a novel CNN algorithm that recognizes the severity of HF and detects early stages of HF with high accuracy, irrespective of EF.

We collected 10-s, 12-lead ECGs from adult patients and healthy individuals, 20 years old or older. We included outpatients with or without cardiovascular disease (CVD) and inpatients with HFrEF or HFpEF. The inpatients were examined and classified using the NYHA classification by expert cardiologists. After screening, 21,292 ECGs from 9,284 participants were deemed eligible for inclusion in the analysis (Figure 1). Excluded (due to missing values) were 4,274 ECGs from 1,419 participants. After preprocessing as described in the Methods, ECG data from lead I were segmented into single heartbeats, and averaged heartbeat waveforms were obtained for each NYHA grade.

**Figure 1.**
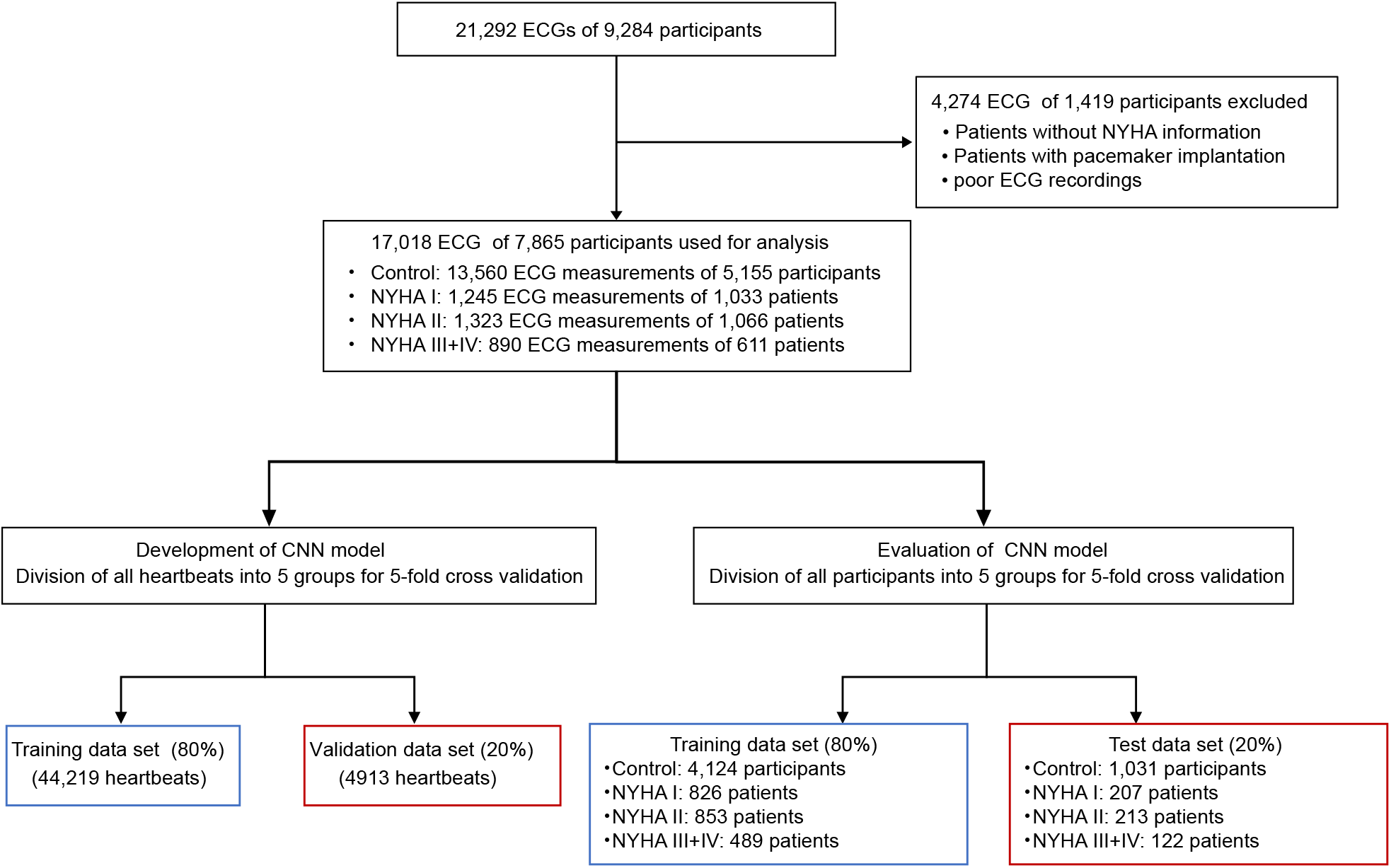
Flow chart for creation of the ECG dataset from HF patients and healthy control. We employed fivefold cross-validation for the development and evaluation of the CNN in two ways. In the first, to develop the CNN the data were randomly split into stratified fivefold subsets of all heartbeat data. The CNN model was trained using 80% of the heartbeat data and optimized using the remaining 20% as an internal validation set. To evaluate the CNN model, the ECG data from the patients were divided into a training dataset and a test dataset. Only heartbeats of patients in the training group were used for development of the CNN; performance of the CNN was then evaluated using only the heartbeats of independent test patients.

We employed fivefold cross-validation for development and evaluation of the CNN (Figure 1). For development of the CNN model, all single beats were spliced from 10-s ECGs from all participants. Among those, 80% were used as beat-by-beat datasets for the training, and the others were used for internal validation. In parallel, the patient-by-patient datasets were divided and used for further CNN training or for the testing data used to evaluate the CNN performance (Figure 1). The patients’ characteristics are shown in Table 1.

**Table 1.** Healthy control and patient characteristics and comorbidities.

|  | Training group | Test group | *P*-value |
| --- | --- | --- | --- |
| Age, years | 59.4 ±14.3 | 58.9±13.7 | 0.07 |
| Female, sex, % | 44.3% | 44.6% | 0.80 |
| Mean EF, % | 56.5±17.3 | 55.5±18.1 | 0.24 |
| History of HF | 34.3% | 34.4% | 0.96 |
| HFpEF | 73.7% | 70.1% | 0.18 |
| HFrEF | 26.3% | 29.9% |  |
| Coronary Artery Disease | 9.9% | 11.2% | 0.18 |

The CNN was designed to use the lead I ECG waveform to distinguish HF from control or to classify the severity of HF, beat by beat, according to the NYHA classification. The CNN included three convolution layers and three max-pooling layers for both training and identification. The learned feature maps were then fed into two fully connected neural networks with an output layer of 4 nodes, which refer to participants without CVD (controls) or patients with NYHA I, II or III-IV HF (Figure 2). The CNN model was able to distinguish the HF patients, including NYHA I, II, III and IV, from healthy controls, with an area under the receiver operating characteristic curve (AUC) of 0.996 (95% CI:0.994-0.997), sensitivity of 96.2% (95% CI: 0.954-0.969), specificity of 98.5% (95% CI: 0.98-0.99), and accuracy of 97.4% (95% CI: 0.969-0.978) (Figure 3a). The CNN model also accurately classified the patients into 3 or 4 groups: control, NYHA I and NYHA II-IV, or control, NYHA I, NYHA II and NYHA III-IV (Figure 3b, 3c and table 2).

**Table 2.** Model performance.

|  |  | AUC | Sensitivity | Specificity | Accuracy |
| --- | --- | --- | --- | --- | --- |
| **2 Groups** | Control | 0.996 (0.994-0.997) | 98.4% (0.977-0.989) | 96.4% (0.956-0.971) | 97.4% (0.969-0.978) |
|  | Heart failure | 0.996 (0.994-0.997) | 96.2% (0.954-0.969) | 98.5% (0.980-0.990) | 97.4% (0.969-0.978) |
| **3 Groups** | Control | 0.993 (0.992-0.995) | 96.7% (0.961-0.974) | 96.0% (0.953-0.966) | 96.3% (0.958-0.967) |
|  | NYHA I | 0.967 (0.963-0.970) | 82.1% (0.802-0.840) | 95.6% (0.950-0.963) | 92.0% (0.914-0.928) |
|  | NYHA II-IV | 0.989 (0.987-0.991) | 92.1% (0.910-0.932) | 96.9% (0.963-0.974) | 95.3% (0.948-0.958) |
| **4 Groups** | Control | 0.995 (0.994-0.996) | 94.8% (0.938-0.956) | 98.1% (0.978-0.984) | 97.2% (0.968-0.975) |
|  | NYHA I | 0.978 (0.974-0.981) | 86.7% (0.855-0.883) | 96.6% (0.961-0.970) | 94.5% (0.939-0.949) |
|  | NYHA II | 0.945 (0.939-0.950) | 79.8% (0.780-0.816) | 93.6% (0.930-0.943) | 89.8% (0.891-0.906) |
|  | NYHA III-IV | 0.965 (0.961-0.969) | 80.0% (0.779-0.819) | 95.9% (0.953-0.963) | 92.7% (0.922-0.933) |

**Figure 2.**
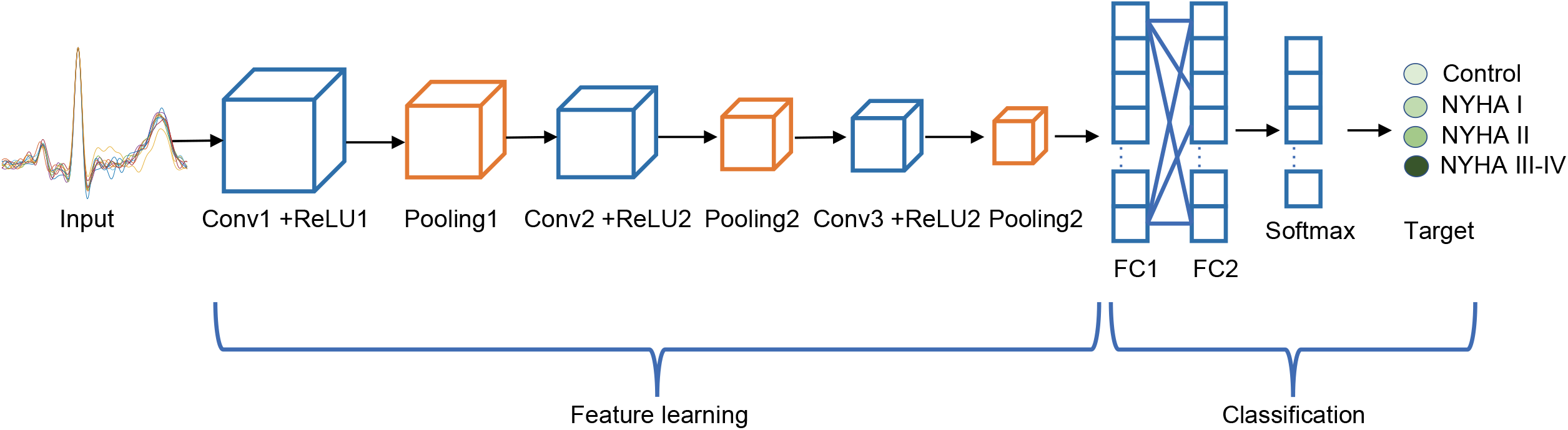
Construction of the neural network architecture for evaluation of ECGs and HF. The CNN included three convolution layers and three max-pooling layers for both the training and identification. We used [1×128], [1×2], and [1×2] as the kernels (filters) to generate the convolutional layers and the max-pooling layers. The learned-feature maps were then fed to two fully connected-neural networks with an output layer of 4 nodes corresponding to control, NYHA I, NYHA II, NYHA III-IV. The back-propagation approach was used to train the CNN.

**Figure 3.**
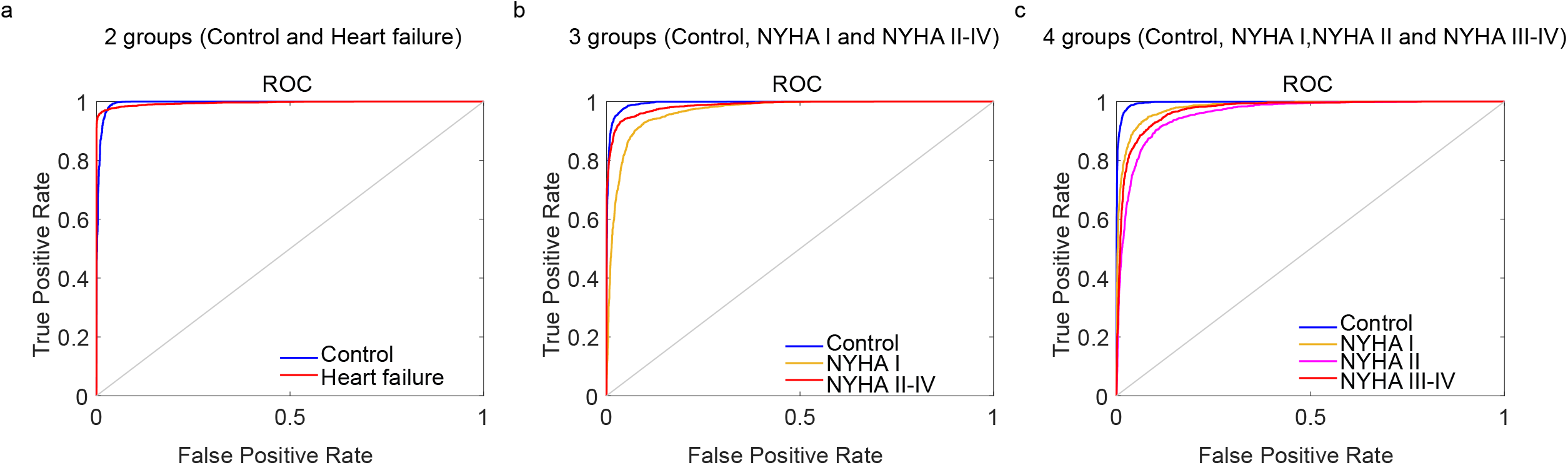
Receiver-operating characteristic (ROC) curves from the convolutional neural network (CNN) for evaluation of HF symptoms with single-lead ECGs. ROC curves for classification with two groups (a), three groups (b), and four groups (c) are shown. The areas under the ROC curves (AUCs) were calculated from the results of performance analyses. High AUCs for all classifications for detection of HF or evaluation of HF severity are shown.

We next investigated which segments of the ECG wave are crucial features of each NYHA classification. We employed Gradient-weighted Class Activation Mapping (Grad-CAM) to highlight the essential regions in the input heartbeats for the classification^15^. The averaged ECG waveform for all heartbeats within each group in the training dataset showed different ECG morphologies (Figure 4). The histograms (gray bars) on the average heartbeat waveforms indicate the essential information transmitted through the network to determine the class-discriminative information in the input time-series.

**Figure 4.**
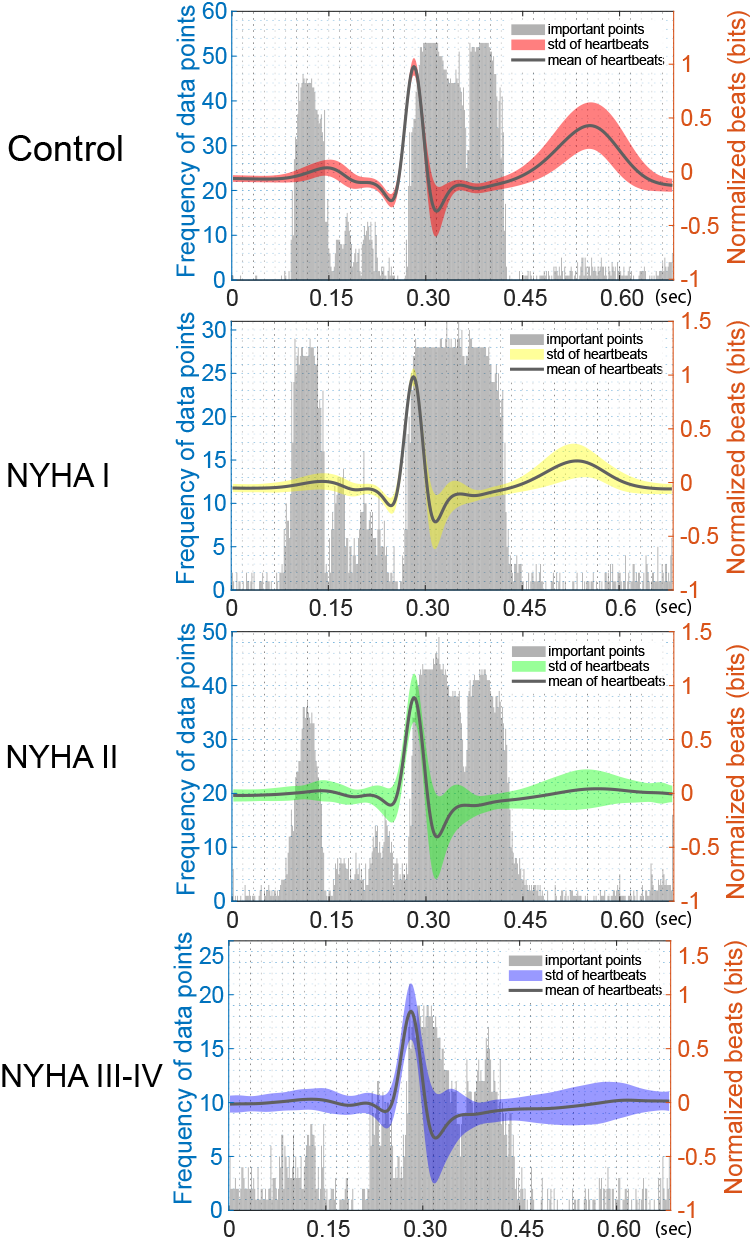
Visualization of feature segments of ECGs used for HF classification by the CNN. The solid lines represent the mean with the respective error bands of all heartbeats that correspond to each classification group in the training datasets. The error bands represent the standard deviation of the heartbeats in each category. The vertical bars represent the histograms of the sample points (=1) obtained by applying Grad-CAM methods to all of the training heartbeats in the normalized heatmaps. The highlighted parts in each ECG waveform represent the ECG heartbeat subsequence used by the CNN to generate a prediction. The histograms of the critical sample points indicate the essential segments of a heartbeat used for classifying groups. Comparison among ECG waveforms from the four classes revealed that the amplitude of the T wave became smaller as HF became more severe. A wide QRS complex was observed in the NYHA III-IV group.In the histogram, the onset of the P wave appears to be important in the Control and NYHA I-II groups, but not the NYHA III-IV group. The beginning of QRS is important in the NYHA III-IV group. The offset of the QRS and the onset of the T wave are highlighted in all groups.

A widened QRS and a prolonged PR interval were previously reported to be independently associated with in-hospital death, post-discharge death, and rehospitalization^16^. However, there is little consensus on the predictive value of ECG characteristics in the HF population^17^. In the present study, comparison among the ECG waveforms in the 4 classes revealed that the severer the HF, the lower the T wave. In addition, a wider QRS complex was observed in NYHA III-IV group. In the Grad-CAM histogram, the onset of the P wave was indicated as an outstanding feature in the control, NYHA I and NYHA II groups, but not the NYHA III-IV group. While the offset of the QRS and the beginning of the T wave were highlighted as important in all groups, the onset of the QRS was marked as especially important in the NYHA III-IV group. These HF class-specific ECG morphologies may provide new insight into how heart burden affects the electrical activity of the heart.

An earlier study reported a CNN that could detect HF using single beats from a small number of patients in a public data base and was able to distinguish HF patients from healthy controls^18^. But while this algorithm could potentially help non-specialists avoid overlooking apparent HF, it is unable to classify the severity of HF. To improve current HF management and avoid its recurrence, detailed classification of symptoms collected from daily life, including mild symptoms, is required.

Our CNN model enables us to distinguish HF patients from healthy controls with high accuracy (AUC 0.996). Moreover, it can provide the severity of HF based on NYHA classification and distinguish between NYHA I and NYHA II, suggesting it could be used for early detection of HF at home. Early detection of HF progression would enable prescription of additional medications before decompensation, which has been linked to decreasing the incidence major adverse cardiovascular events and unplanned hospital admissions19. Our study is the first to report a CNN able to classify the severity of HF. It is expected that applying our AI algorithm to mobile single-lead ECG recorders will enable patients themselves to detect HF early through self-monitoring.

Levels of biomarkers such as BNP and NT-pro-BNP are often used when diagnosing or monitoring HF^20^, and reductions in these biomarkers may predict a better prognosis^21^. However, some clinical trials have not found a benefit in terms of quality of life or cardiovascular outcomes of even frequent monitoring of these biomarkers to assess whether patients need additional treatment for their HF^22-24^.

Continuous monitoring of heart rate, heart rate variability, accelerometry, respiratory rate, and body temperature with a wearable multisensory device is reportedly able to detect impending rehospitalization of HF patients with predictive accuracy (AUC of 0.88)^25^. The accuracy of our CNN model (AUC 0.996) for detection of HF is superior to the medical screening test based on biomedical sensors or BNP (AUC 0.60)^24^.

This study shows the feasibility of using CNN models of single lead ECG data to improve diagnosis of HF severity, irrespective of EF. Since the lead I ECG can be easily collected during daily life by using a portable or wearable ECG monitor, our algorithm may help with remote management of HFrEF and HFpEF.

## Data Availability

No additional data is available.

## Methods

### Data sources and study population

All data used in this study were acquired between 2013 and 2019 at the University of Tokyo hospital. This study was approved by the institutional ethical committee of the University of Tokyo (No. 2020024NI-(1)). All participants were at least 20 years old. The control group was composed of healthy individuals who underwent a comprehensive medical examination and were diagnosed as having no illness. All patients with HF were diagnosed as New York Heart Association (NYHA) I, NYHA II, NYHA III, or NYHA IV according to NYHA functional classification criteria^26^. The definitions of the classifications are as follows:

1. Control: Participants have no cardiovascular heart disease (CVD).
2. NYHA I: Patient has been hospitalized for HF but has no limitation of physical activity; physical activity does not cause fatigue, palpitation or dyspnea.
3. NYHA II: Patient has been hospitalized for HF, and physical activity is slightly limited by fatigue, palpitation or dyspnea, but they have no symptoms at rest.
4. NYHA III: Patient has been hospitalized for HF, and physical activity is greatly limited by fatigue, palpitation or dyspnea, even at rest.
5. NYHA IV: Patient has been hospitalized for HF and cannot carry on any physical activity without discomfort, and they experience symptoms of HF at rest.

ECG data were obtained from the control group at an annual health check center at the University of Tokyo hospital, where individuals receive regular yearly health checks. ECG data were obtained from patients clinically diagnosed with HF during their hospitalization in the department of cardiology. Standard 10-s, 12-lead ECGs were recorded in all patients with HF on admission, and a NYHA class was assigned based on the patient’s symptoms and evaluation of their medical examination by expert cardiologists. Data lacking a NYHA classification were excluded, as were data from patients with a pacemaker and poor recordings due to motion artifacts, inaccurate electrode applications and excessive noise. Used in the study were 17,018 10-s ECGs from 7,865 patients.

### CNN model development

Standard 10-s, 12-lead ECGs were recorded at 500 Hz with patients in a supine position in a resting state. Only lead-I ECG data were selected for this study. We segmented the 10-s ECG recordings into heartbeat waveforms as independent input data for training of the CNN model^27^. Before heartbeat segmentation, preprocessing of ECG recordings was conducted to eliminate baseline drift and noise. First, the baseline drift was removed using the wavelet decomposition method. ECG data were decomposed into sublevels, and the final approximation coefficient was taken as the baseline drift and subtracted from the original signal. Next, a Butterworth bandpass filter was applied to remove power-line noise and high-frequency distortion. Thereafter, heartbeats were detected by using the Pan-Tompkins algorithm to recognize the peaks of the R waves on the ECG recordings^28^. A window was then used to segment the heartbeats between 0.28 s before and 0.40 s after the R-wave peaks to capture the PQRST complexes. This process enabled us to adjust the alignment of heartbeats using R-wave peaks. After preprocessing, all segmented heartbeats were normalized to between −1 and 1.

Heartbeats were annotated based on the NYHA classification of corresponding 10-s ECG recordings. To evaluate the generalizability and stability of the proposed algorithm, we combined two or more NHYA classes into one group. We then trained and tested the model with a two-group dataset, three-group dataset, and four-group dataset, separately. Two groups refers to Control vs. the patients with HF. Three groups refer to Control, NYHA I and NYHA II-IV. NYHA II-IV was further divided into NYHA II and NYHA III-IV and used as four-group dataset.

To increase computational efficiency, we applied vector quantization (VQ) for data sampling. The VQ method^29^ is an efficient technique for data compression, which can map the original data into prototype vectors to avoid loose information as much as possible. The VQ process included two steps: designing a representative codebook, then assigning a label to each input datum (heartbeat) from the codebook. By labeling all heartbeats according to the designed codebook, we were able to divide heartbeats into clusters. The number of clusters is the size of the codebook. For each cluster, the frequency distribution of heartbeats in each NYHA class was determined. We removed heartbeats with a low frequency (less than 25%) from NYHA classes and retained the remaining heartbeats as representative of the corresponding class for training and validation. To achieve an “optimal” codebook size, we varied the size from 4 to 256 and evaluated the classification accuracy. The best performances were observed with codebook sizes of 8, 16, and 16 for two groups, three groups, and four groups, respectively.

### CNN modeling and validation

The learned features reflected by the CNN parameters were then used to identify the NYHA class from the test heartbeat waveform. The CNN takes heartbeat waveforms as one-dimensional time-series inputs and outputs label predictions as NYHA classes. In our study, the CNN model included three convolution layers and three max-pooling layers for both training and identification. Each convolutional layer was followed by batch normalization, rectified linear unit (ReLU) activation, and a 10% dropout to regularize many parameters. To be specific, the kernels (filters) used to generate the convolutional layers were chosen as [1×128], [1×2], and [1×2], respectively. The learned feature maps were then fed into two flattened and fully connected layers followed by a softmax layer with an output layer of nodes corresponding to the NYHA classes.

The proposed model was trained and validated using fivefold cross-validation, whereby the data were randomly split into stratified fivefold subsets. The CNN model was trained using 80% of the heartbeats and evaluated using the remaining 20%. Thereafter, 20% of the data were randomly selected as an internal validation set. Overfitting was avoided by tracking the validation loss during training. This process was repeated five times to obtain a mean classification rate. During the cross-validation process, each subset of heartbeats was taken for internal validation, enabling an unbiased estimate of the classification error rate.

The fivefold cross-validation was performed in two patterns. In the first, all heartbeats were randomly divided into training and test sets, irrespective of the patients. This pattern makes each heartbeat an independent input, which enables an efficient global evaluation. However, there is the possibility that heartbeats from the same 10-s ECG recording are simultaneously included in both the training and test set. In the second pattern, all patients were divided into a training and a test group to avoid overlap of patients between the training and test sets. Only heartbeats from patients assigned to the training dataset were used for modeling. The performance was then evaluated solely based on the heartbeats of the independent test dataset.

### Visual explanation of the CNN model used to identify heart failure and NYHA classification

To understand which parts of the heartbeat waveform are most important for its NYHA classification, gradient-weighted class activation mapping (Grad-CAM) was used to show the gradient of the classification score for the convolutional features determined by the network. This can help a clinician understand why the CNN model makes a given classification. The idea of Grad-CAM is to calculate the gradient of the final classification score for the final convolutional feature map. The places where this gradient is large are where the final score depends mostly on the data. In other words, the data points on the heartbeat waveform that have the highest Grad-CAM scores contribute most to the classification. Specifically, to create an ‘average’ Grad-CAM for each NYHA class, we calculated the respective pointwise Grad-CAM scores (normalized to between 0 and 1) for each heartbeat. We then calculated the average heartbeat waveform for each class and accumulated the occurrence of data points that had Grad-CAM scores equal to 1. From the obtained ‘average’ heartbeat waveform and the frequency map, the places where high Grad-CAM scores most frequently occur indicate significant features that have the most impact on the classification.

### Statistical considerations

We compared baseline demographics between patients. Statistical significance was tested between the groups using a chi-square test for categorical variables. The other categorical variables are presented as numbers (percent) and were compared by using unpaired Student’s t-tests. All analyses were two-sided, and values of *P* less than 0.05 were considered statistically significant. □ To quantify the validation performance, we estimated the area under the curve (AUC) and the sensitivity, specificity, and accuracy with 95% confidential intervals from the ROC curves. Accuracy was evaluated for the optimal operating point on the ROC curve that maximized the sensitivity and specificity. We averaged the AUC, sensitivity, specificity, and accuracy of the fivefold subsets to incorporate the uncertainty when selecting cutoff points. The 95% CI was estimated by bootstrapping 1,000 random and variable sampled instances. All training and validation were implemented using MATLAB R2020a and performed on an NVIDIA GeForce RTX 2080 Ti platform.

## Author contributions

E.H. and K.F. contributed to study design, study coordination, analyzed the data, data interpretation, and wrote the manuscript. E.H. and Y.S. contributed to data collection and data management. T.O., T.M., N.F., J.M., L.Y., J.S., Y.N., H.M., A.S., Y.M., T.O. and G.O. contributed to analyzing the ECG interpretation. Y.C. contributed to the data analysis and writing the manuscript. M.O. and I.K was the senior supervisor of the project. All author takes responsibility for all aspects of the reliability and freedom from bias of the data presented and their discussed interpretation.

## Acknowledgments

None.

## Corresponding author

Correspondence to Katsuhito Fujiu or Issei Komuro

## References

1. Attia, Z.I., et al. Screening for cardiac contractile dysfunction using an artificial intelligence-enabled electrocardiogram. Nat Med 25, 70–74 (2019).

2. Global, regional, and national incidence, prevalence, and years lived with disability for 354 diseases and injuries for 195 countries and territories, 1990-2017: a systematic analysis for the Global Burden of Disease Study 2017. Lancet 392, 1789-1858 (2018).

3. Groenewegen, A., Rutten, F.H., Mosterd, A. & Hoes, A.W. Epidemiology of heart failure. Eur J Heart Fail (2020).

4. Ponikowski, P., et al. 2016 ESC Guidelines for the diagnosis and treatment of acute and chronic heart failure: The Task Force for the diagnosis and treatment of acute and chronic heart failure of the European Society of Cardiology (ESC). Developed with the special contribution of the Heart Failure Association (HFA) of the ESC. Eur J Heart Fail 18, 891–975 (2016).

5. Cho, J., et al. Artificial Intelligence Algorithm for Screening Heart Failure with Reduced Ejection Fraction Using Electrocardiography. Asaio j (2020).

6. Triposkiadis, F., et al. The continuous heart failure spectrum: moving beyond an ejection fraction classification. Eur Heart J 40, 2155–2163 (2019).

7. Solomon, S.D., et al. Influence of Ejection Fraction on Cardiovascular Outcomes in a Broad Spectrum of Heart Failure Patients. Circulation 112, 3738–3744 (2005).

8. Metra, M. & Teerlink, J.R. Heart failure. The Lancet 390, 1981–1995 (2017).

9. Steinberg, B.A., et al. Trends in patients hospitalized with heart failure and preserved left ventricular ejection fraction: prevalence, therapies, and outcomes. Circulation 126, 65–75 (2012).

10. Dunlay, S.M., Roger, V.L. & Redfield, M.M. Epidemiology of heart failure with preserved ejection fraction. Nature Reviews Cardiology 14, 591–602 (2017).

11. Flachskampf, F.A., et al. Cardiac Imaging to Evaluate Left Ventricular Diastolic Function. JACC Cardiovasc Imaging 8, 1071–1093 (2015).

12. Ammar, K.A., et al. Prevalence and prognostic significance of heart failure stages: application of the American College of Cardiology/American Heart Association heart failure staging criteria in the community. Circulation 115, 1563–1570 (2007).

13. Dickstein, K., et al. ESC guidelines for the diagnosis and treatment of acute and chronic heart failure 2008: the Task Force for the diagnosis and treatment of acute and chronic heart failure 2008 of the European Society of Cardiology. Developed in collaboration with the Heart Failure Association of the ESC (HFA) and endorsed by the European Society of Intensive Care Medicine (ESICM). Eur J Heart Fail 10, 933–989 (2008).

14. Pascual-Figal, D. & Bayes-Genis, A. The misperception of ‘stable’ heart failure. Eur J Heart Fail 20, 1375–1378 (2018).

15. Selvaraju, R.R., et al. Grad-CAM: Visual Explanations from Deep Networks via Gradient-Based Localization. International Journal of Computer Vision 128, 336–359 (2020).

16. Park, S.-J., et al. Short- and long-term outcomes depending on electrical dyssynchrony markers in patients presenting with acute heart failure: Clinical implication of the first-degree atrioventricular block and QRS prolongation from the Korean Heart Failure registry. American Heart Journal 165, 57-64.e52 (2013).

17. Gouda, P., Brown, P., Rowe, B.H., McAlister, P. & Ezekowitz, J.A. Insights into the importance of the electrocardiogram in patients with acute heart failure. European Journal of Heart Failure 18, 1032–1040 (2016).

18. Porumb, M., Iadanza, E., Massaro, S. & Pecchia, L. A convolutional neural network approach to detect congestive heart failure. Biomedical Signal Processing and Control 55, 101597 (2020).

19. Halliday, B.P., et al. Withdrawal of pharmacological treatment for heart failure in patients with recovered dilated cardiomyopathy (TRED-HF): an open-label, pilot, randomised trial. The Lancet 393, 61–73 (2019).

20. Members, A.T.F., et al. ESC Guidelines for the diagnosis and treatment of acute and chronic heart failure 2012: The Task Force for the Diagnosis and Treatment of Acute and Chronic Heart Failure 2012 of the European Society of Cardiology. Developed in collaboration with the Heart Failure Association (HFA) of the ESC. European Heart Journal 33, 1787–1847 (2012).

21. Masson, S., et al. Prognostic value of changes in N-terminal pro-brain natriuretic peptide in Val-HeFT (Valsartan Heart Failure Trial). J Am Coll Cardiol 52, 997–1003 (2008).

22. Pfisterer, M., et al. BNP-guided vs symptom-guided heart failure therapy: the Trial of Intensified vs Standard Medical Therapy in Elderly Patients With Congestive Heart Failure (TIME-CHF) randomized trial. Jama 301, 383–392 (2009).

23. Eurlings, L.W., et al. Management of chronic heart failure guided by individual N-terminal pro-B-type natriuretic peptide targets: results of the PRIMA (Can PRo-brain-natriuretic peptide guided therapy of chronic heart failure IMprove heart fAilure morbidity and mortality?) study. J Am Coll Cardiol 56, 2090–2100 (2010).

24. Bhalla, V., et al. Diagnostic ability of B-type natriuretic peptide and impedance cardiography: testing to identify left ventricular dysfunction in hypertensive patients. Am J Hypertens 18, 73s–81s (2005).

25. Stehlik, J., et al. CONTINUOUS WEARABLE MONITORING ANALYTICS PREDICT HEART FAILURE DECOMPENSATION: THE LINK-HF MULTI-CENTER STUDY. Journal of the American College of Cardiology 71, A646 (2018).

26. Dolgin, M. & Committee, N.Y.H.A.C. Nomenclature and criteria for diagnosis of diseases of the heart and great vessels, (Little, Brown, 1994).

27. Shin, H.-C., et al. Deep Convolutional Neural Networks for Computer-Aided Detection: CNN Architectures, Dataset Characteristics and Transfer Learning. IEEE Trans Med Imaging 35, 1285–1298 (2016).

28. Pan, J. & Tompkins, W.J. A Real-Time QRS Detection Algorithm. IEEE Transactions on Biomedical Engineering BME-32, 230–236 (1985).

29. Gersho, A. & Gray, R.M. Vector Quantization and Signal Compression, (Springer US, 1991).

